# Treadmill training with rhythmic auditory cueing and/or visual feedback for persons with Multiple Sclerosis: feasibility and effects on gait parameters in a clinical randomized controlled trial

**DOI:** 10.64898/2026.06.18.26356023

**Authors:** Philipp Kröber, Florian Wolf, Jochen Saliger, Jörn Nielsen, Mareike Eschweiler

## Abstract

**Background:** Gait training incorporating visual feedback or rhythmic auditory cueing has shown promising results in neurological conditions but has rarely been investigated in clinical rehabilitation for persons with Multiple Sclerosis (pwMS).

**Objective:** To evaluate the feasibility of treadmill training (TT) with visual feedback (VF) and TT with visual feedback plus rhythmic auditory cueing (VF+RAC) during clinical rehabilitation and explore its effects on gait parameters.

**Methods:** PwMS were randomly allocated 1:1 to perform ten 30-minute training sessions of TT with VF or VF+RAC during inpatient rehabilitation. The primary outcome was feasibility (adherence, compliance, safety, and acceptability). Secondary outcomes were session-by-session developments in spatiotemporal and qualitative gait parameters.

**Results:** Sixty of 68 randomized participants completed the intervention (VF: n=29; VF+RAC: n=31). Adherence and compliance rates were 93% and 86%, respectively, with no differences between groups. The most common adverse event in both groups was (leg) pain (21/38 total adverse events). One fall occurred in 629 sessions. Both interventions were greatly accepted and perceived as fun, motivating and helpful to achieve rehabilitation goals. Both groups increased in distance, gait speed, and average step length. Step length variability did not change in the VF-group, while the VF+RAC-group slightly improved. Step length difference was constantly low in the VF+RAC-group, while the VF-group differences were elevated.

**Conclusions:** VF and VF+RAC are feasible training options for pwMS in a rehabilitation setting and are greatly accepted by participants. Qualitative gait parameters should be investigated in studies powered to detect clinically relevant differences in the future.

**Trial registration:** German Clinical Trials Register: DRKS00023709, date of registration: 07 January 2021

## 1 Introduction

Gait impairment is one of the most prominent symptoms of Multiple Sclerosis, with 41% of persons with MS (pwMS) reporting difficulties while walking, of which 70% describe it as the most challenging aspect of living with MS (LaRocca, 2011). Compared to healthy adults, pwMS take smaller steps and walk at a slower pace (Coca-Tapia et al., 2021; Socie et al., 2013), have worse walking endurance (Cederberg et al., 2019), and exhibit larger step length variability and step asymmetry (Kalron, 2015; Socie et al., 2013; Timmermans et al., 2025). To compensate for these gait deficits and maintain a safe gait and postural control, utilization of cognitive resources is necessary (Santinelli et al., 2021). This makes walking a motor-cognitive dual-task that requires more cognitive resources through prefrontal cortex activation than for healthy people (De Sanctis et al., 2020; Kim and Fraser, 2022), potentially contributing to perceived fatigue and walking fatigability in pwMS (Santinelli et al., 2024).

Through technological advancements, new multi-component interventions such as virtual reality, walking with feedback, or walking with rhythmic auditory cueing (RAC) have shown promising results to increase functioning in pwMS, especially walking ability (Galperin et al., 2023; Maggio et al., 2021; Seebacher et al., 2024).

In RAC, a metronome or music provides a continuous beat, while footsteps must be synchronized with the beat (‘auditory-motor coupling’) (Ziane and Dalla Bella, 2026). PwMS are able to achieve synchronization but this is influenced by cognitive flexibility, working memory, and dynamic balance (Vanbilsen et al., 2026, 2024). First studies confirmed the benefits of RAC on walking through a reduction of activation in motor planning and attention areas in the brain, suggesting more efficient recruitment of areas subserving motor function (Helmlinger et al., 2025). Improved spatiotemporal gait parameters by applying RAC comprise step length, cadence and gait speed and incites to explore the effects on larger samples of pwMS in clinical rehabilitation settings (Ghai and Ghai, 2018). Although qualitative gait parameters such as step length variability and step length symmetry are also paramount for normal gait, energy expenditure, and perceived effort while walking (Baček et al., 2025; Ellis et al., 2013; Kalron, 2015; Socie et al., 2013; Timmermans et al., 2025), no study has investigated the effects of RAC on these parameters in pwMS.

Another approach of motor-cognitive walking training utilizes visual feedback. Participants receive live feedback on gait parameters and are encouraged to adapt their gait to the feedback. Unlike RAC, where cues are given at a predetermined regularity, independent of the patient’s actions, feedback is based on the patient’s previous steps. Therefore, adaptation to feedback relies on the patient’s ability to analyze, process and adapt their gait pattern based on the information given. Visual feedback has been shown to be a feasible training tool in persons with neurological conditions, including stoke, Parkinson’s disease and mixed conditions and can improve gait parameters such as step length and gait speed (Gooßes et al., 2020; Kaźmierczak et al., 2022; Kim and Oh, 2020; Silva-Batista et al., 2023). However, to the best of our knowledge, interventions using visual feedback with pwMS have not been conducted yet.

One small exploratory study including ten persons with secondary progressive MS in the experimental group, evaluated an intervention using a treadmill that can provide a combination of visual feedback and RAC at the same time, which showed promising results regarding selected aspects of feasibility and motor outcomes (Maggio et al., 2021).

For clinical practice, no data exists regarding which exercise duration and frequency is acceptable and safe for pwMS when performing walking training with RAC and/or visual feedback. This is critical information, necessary for further intervention development (Declerck et al., 2025; Motl et al., 2023).

Therefore, we aimed to investigate whether adding treadmill training with visual feedback or visual feedback plus RAC to the rehabilitation schedule of pwMS in a German inpatient rehabilitation clinic, is feasible regarding participant adherence, compliance, safety, and acceptability. Furthermore, we aimed to explore the effects of visual feedback and visual feedback plus RAC on spatiotemporal and qualitative gait parameters.

## 2 Methods

### 2.1 Design and setting

This stage Ib feasibility study (Declerck et al., 2025) was located at the Johanniter-Klinik Godeshöhe (JKG) GmbH in Bonn, Germany, which is a neurological rehabilitation center. The study had a two-armed, parallel group, randomized-controlled design with a 1:1 allocation ratio. We intended to recruit 34 participants per group. Feasibility data were collected throughout the intervention period and post-intervention, while gait parameters were collected for each session by the Gait Trainer 3.1 treadmill (Biodex Medical System, 20 Ramsay Road, Shirley, NY, USA). We also performed clinical motor and cognitive assessments and administered patient-reported outcomes, which will be reported separately. All sessions on the Gait Trainer 3.1 were scheduled additionally to the participant’s rehabilitation therapies, which they received based on individual needs.

Ethical approval was obtained from the Ethics Committee of the medical faculty of the University of Bonn (reference number: 535/20). The study was prospectively registered in the German Clinical Trials Register (ID: DRKS00023709, date of registration: 07 January 2021). Reporting of this trial follows the CONSORT reporting guidelines for randomized pilot and feasibility trials (Eldridge et al., 2016) (Supplement 1).

### 2.2 Participants

Admissions were screened for pwMS and eligibility criteria evaluated by neuropsychologists (JS and JN). If deemed eligible, pwMS were informed about the study verbally and in written form. Inclusion criteria were confirmed diagnosis of MS based on the 2017 McDonald criteria (Thompson et al., 2018), no contraindications regarding treadmill training (e.g., severe orthopedic or cardiovascular disorders), a scheduled rehabilitation period adequate to complete all study-related assessments and training sessions (4 weeks), no other neurological or psychiatric disorders, impacting the ability to understand the assessment and training regimen, exercise safety, or additionally impacting motor or cognitive function. All participants had to be able to provide written informed consent. Exclusion criteria consisted of insufficient German language skills, uncorrected visual, or hearing function, severe cognitive deficits (indicated by neuropsychological testing below the fifth percentile in at least three of five domains of cognitive function: visuospatial abilities, attention, orientation, learning/memory, and executive function), and participation in another exercise-based study.

### 2.3 Randomization and blinding

After providing written informed consent, participants were randomly allocated to treadmill training with visual feedback (VF) or treadmill training with visual feedback plus rhythmic auditory cueing (VF+RAC), without stratification. Using the https://randomizer.org platform, study IDs were randomized before commencement of recruitment by ME. Group assignment during the study period was performed by PK, JS and ME.

Since randomization was performed after the pre-intervention assessments, group allocation was concealed for participants and study staff at that point. Due to the study design and resources available, further blinding could not be upheld for training interventions and post-assessments.

### 2.4 Interventions

The intervention period comprised ten 30-minute training sessions, which were usually conducted within a 2-week period on weekdays. Both groups exercised on the Gait Trainer 3.1 treadmill, which can provide visual feedback and RAC. A timeline for the training sessions is displayed in Figure 1.

**Figure 1.**
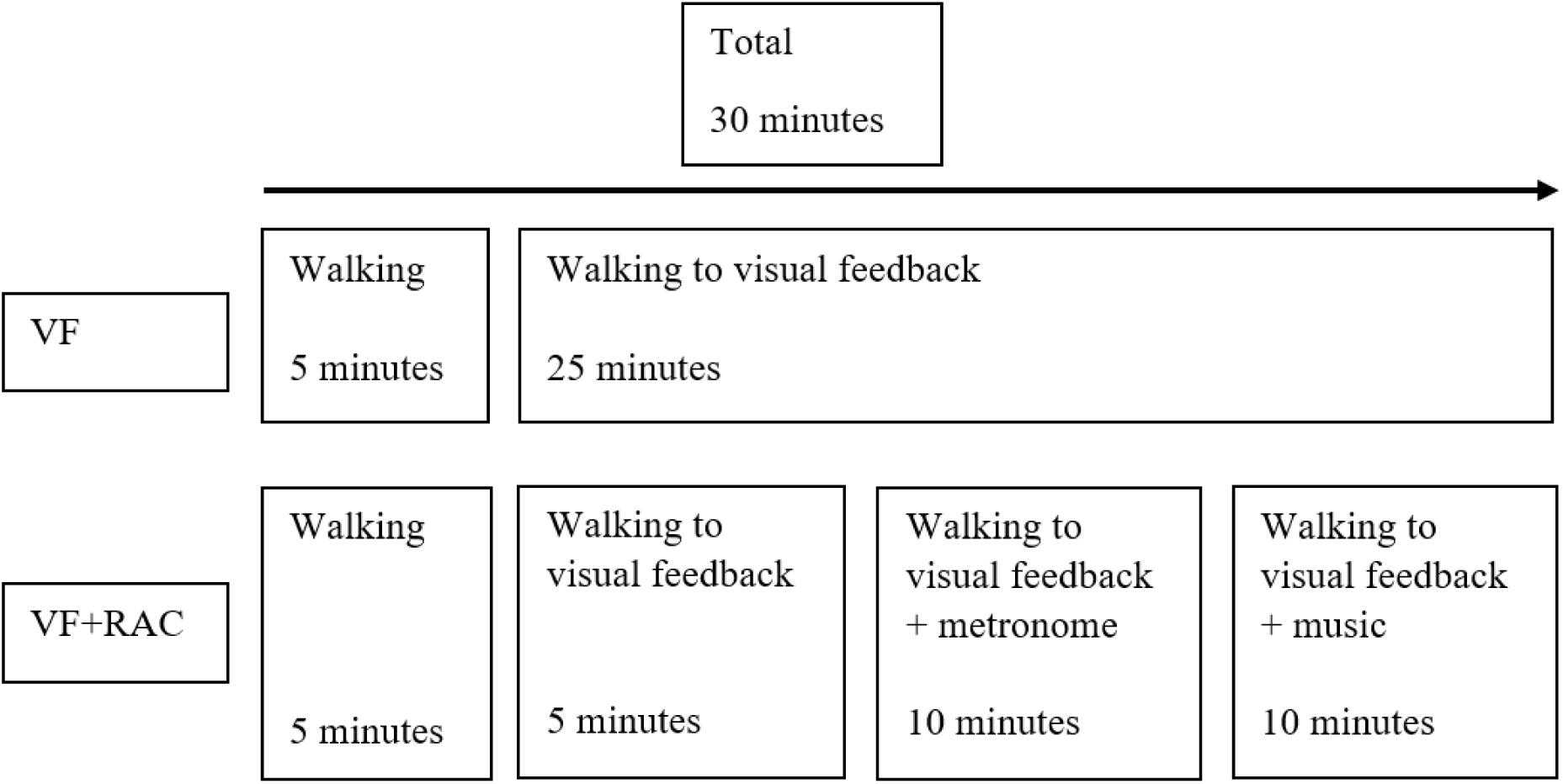
Schedule for training sessions. VF=Visual feedback; VF+RAC=Visual feedback+rhythmic auditory cueing

The initial 5 minutes of the training sessions were identical for both groups and were spent walking on the treadmill without any additional task. Initial speed of the treadmill was determined by the cadence of the participant’s usual overground walking, evaluated during the pre-intervention assessment.

After 5 minutes, a visual feedback was presented to participants of both groups on a screen, which was affixed to the treadmill (Figure 2). The feedback displayed the participant’s average step length of the last five steps as measured by sensors underneath the treadmill belt. Additionally, it indicated the optimal range for the participant’s individual step length, and provided feedback whether the step length was excessive, insufficient, or within the optimal range. The optimal range is calculated by the Gait Trainer 3.1 based on normative values (Whittle, 1997), according to the participant’s height, age, and gender. The visual feedback remained visible until the end of the session for both groups. For the VF-group no further changes were made until the end of the session.

**Figure 2.**
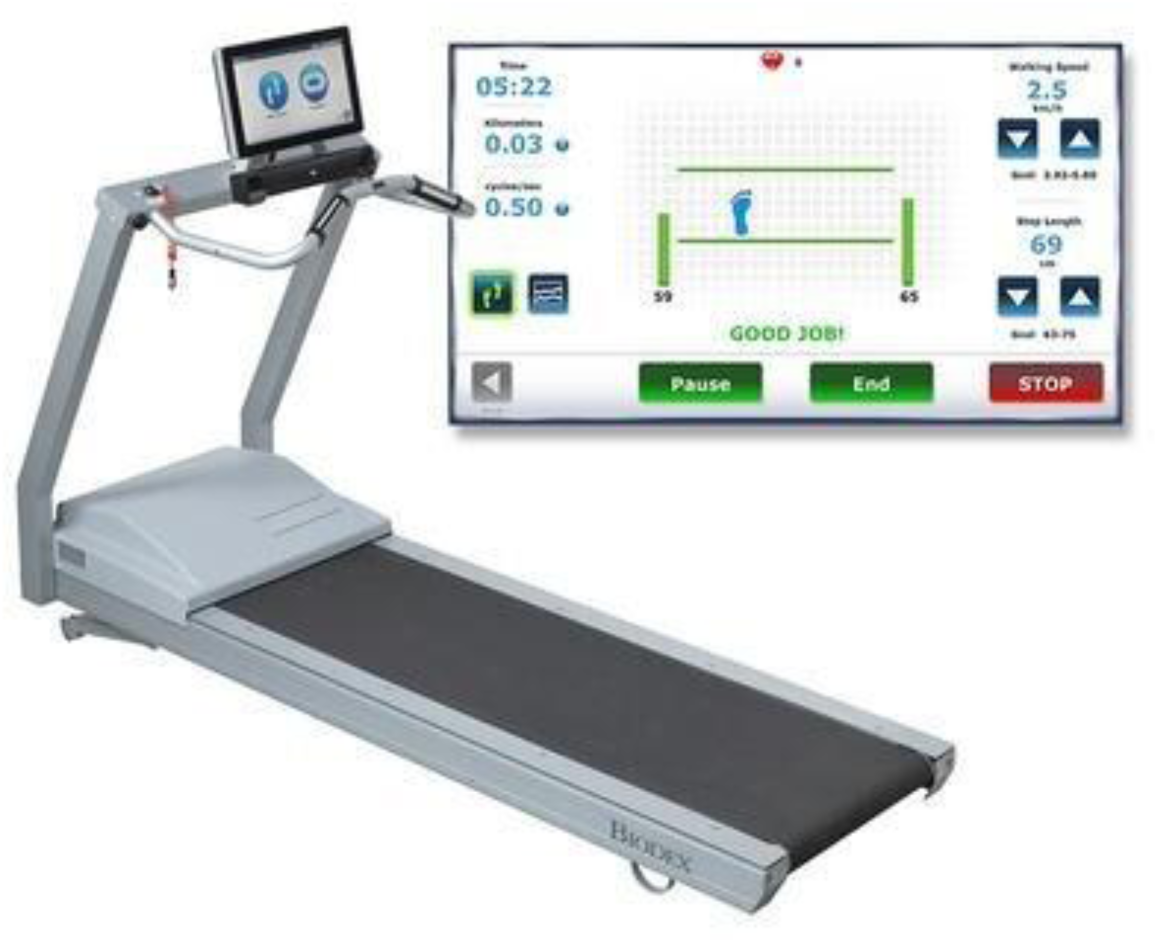
Gait Trainer 3.1 including visual feedback screen.

Ten minutes into the session, a metronome was added only for the VF+RAC-group. The metronome’s rhythm was calibrated to align with the optimal cadence for the participant (e.g., for 100 steps/min, the metronome was set to 100 bpm), whose task was to synchronize their steps to the metronome, i.e. place one foot to every beat of the metronome. After 20 minutes, the metronome was replaced by instrumental music created specifically for the Gait Trainer 3.1 by professional music therapists. All participants in the VF+RAC-group exercised to the same piece (‘Animals Everywhere’), which comprises a variety of instruments, introduced sequentially while the underlying rhythm remains audible. The speed of the music was again aligned with the optimal cadence for the individual participant and again, participants were prompted to place their feet to the beat of the music. To ensure an optimal level of challenge, the speed of the treadmill was always adaptable, but did not follow a predetermined schedule. When participants were not able to synchronize their steps to the beat of the metronome or music, the metronome’s/music’s tempo could be adjusted up to ten percent (slower or faster).

During all training sessions there was always the possibility to take rest breaks.

### 2.5 Outcomes

#### 2.5.1 Sociodemographic and clinical data

The participant’s age, sex, MS subtype, time since MS diagnosis, Expanded Disability Status Scale (EDSS), and MS-medication were taken from medical records. The Multiple Sclerosis Functional Composite was calculated using the Paced Auditory Serial Addition Test and the Nine-Hole-Peg Test, which are standard assessments for pwMS in the JKG, as well as the Timed 25-Foot Walk Test, that was part of the baseline motor assessment. Education and musicality were queried. Further, we monitored each participant’s total time spent in rehabilitation activities (i.e., study-related and study-unrelated) that had either a motor or cognitive focus to evaluate whether both groups received similar total amounts of rehabilitation services in these areas during the intervention period.

#### 2.5.2 Feasibility

Adherence was calculated as the number and proportion of training sessions that were conducted compared to all sessions that were planned (i.e., ten for each participant). Furthermore, the number and proportion of participants attending a pre-specified minimum number of eight sessions was analyzed. This was set as the minimum number for inclusion in data analysis for motor and cognitive outcomes, which will be reported elsewhere. Dropouts were defined as participants who withdrew from intervention participation, did not complete at least eight trainings or did not complete the post-intervention assessments.

Compliance was calculated as the number and proportion of sessions completed with the full duration of 30 minutes on the treadmill, relative to the number of sessions planned. Moreover, the time after which preliminarily terminated trainings ended and the number and time of rest breaks during the trainings were monitored. The ability to synchronize steps to the metronome and music was also evaluated as part of compliance via video recordings of the RAC sessions. 100% synchrony was achieved when a foot was placed (heel strike) to every beat of the metronome/music.

Safety was assessed through tracking of adverse events (AEs) and serious adverse events (SAEs). AEs were defined according to published criteria, as unfavorable outcomes that were noticed during or after the intervention, and SAEs as unfavorable outcomes that result in death or are life-threatening, requiring hospitalization or resulting in permanent disability that occurs during or after the intervention, respectively (Learmonth et al., 2023). If an AE occurred during a session (e.g., leg pain), participants decided whether they were able to continue. Therefore, multiple AEs could be recorded within one session, e.g., pain and a fall.

Four different aspects of acceptability were assessed. First, participants listed their rehabilitation goals, pre-intervention. Post-intervention, they rated how the training contributed to achieve their goals on a 4-point Likert scale (‘not at all relevant’, ‘less relevant’, ‘rather relevant’, and ‘highly relevant’). Second, system usability of the Gait Trainer 3.1 was assessed by administering the System Usability Scale (SUS) (Brooke, 1996), post-intervention. It consists of 10 statements with a 5-point Likert scale (‘do not agree at all’ to ‘fully agree’). Its score ranges from 0 to 100 with higher scores indicating better usability. Third, the treadmill training itself was evaluated post-intervention, by utilizing an adapted version of the Training Evaluation Index (TEI) in an interview format (Ritzmann et al., 2014). Thirteen questions assess aspects such as subjective enjoyment and perceived difficulty, which can be answered with ‘yes’ or ‘no’, totaling to a maximum of 13 points (i.e., best experience with the training). Fourth, before every training, participants rated their current mood and their motivation for the training. After the session, they rated how exhausted they felt and how much fun the training session was. For these queries, 4-point Likert scales were utilized (Supplement 2).

#### 2.5.3 Gait parameters

The treadmill automatically recorded the following gait parameters for each session via its built-in force platform: distance walked, average gait speed, average step length (left and right side) and a coefficient of variation (CoV) for the left and right side. The CoV indicates the variability of step length. Additionally, we calculated the average difference in step length between the left and right leg, i.e., a parameter of gait symmetry.

### 2.6 Data analysis

To determine differences between groups at baseline and session 1 for gait parameters, normally distributed, continuous variables were compared with the independent samples *t*-test, ordinal or non-normally distributed variables were analyzed with the Mann-Whitney-U test, and count data were examined by utilizing the Chi-squared or Fisher’s exact test.

Feasibility data for adherence, compliance, and safety were analyzed descriptively and risk ratios for the occurrence of AEs in individual participants and for the occurrence of AEs in trainings were calculated.

For acceptability, rehabilitation goals were categorized by PK (see Supplement 2.1) and differences in perceived goal achievement were determined by Fisher’s Exact Test. The SUS and TEI were compared with the Mann-Whitney-U Test. Emotions before (mood, motivation) and after training (exhaustion, fun) were analyzed descriptively as well.

Due to lack of power and *a priori* description of meaningful treatment effects on treadmill-based gait parameters, no inferential analysis was performed (Sim, 2019). Instead, we focused on visual inspection of the session-by-session results and mean change scores (session 10 – session 1) with 95% confidence intervals (*CI*s).

Level of significance was 5% for all statistical tests. All analyses were per protocol and performed using R (Version 4.5.2) (R Core Team, 2021).

## 3 Results

### 3.1 Participants

The participant flow is depicted in the CONSORT diagram (Hopewell et al., 2025) (Figure 3). From January 14, 2021 to November 8, 2024, 68 pwMS were recruited and randomized to VF (*n*=34) and VF+RAC (*n*=34). A total of 438 pwMS were admitted to the JKG in this period. However, eligibility criteria were not monitored for all admissions. Factors contributing to discontinuous screening/enrolment were participation in a different exercise study that took place at the study center from November 2021 to November 2022, measures against Covid-19, and limited availability of the study staff.

**Figure 3.**
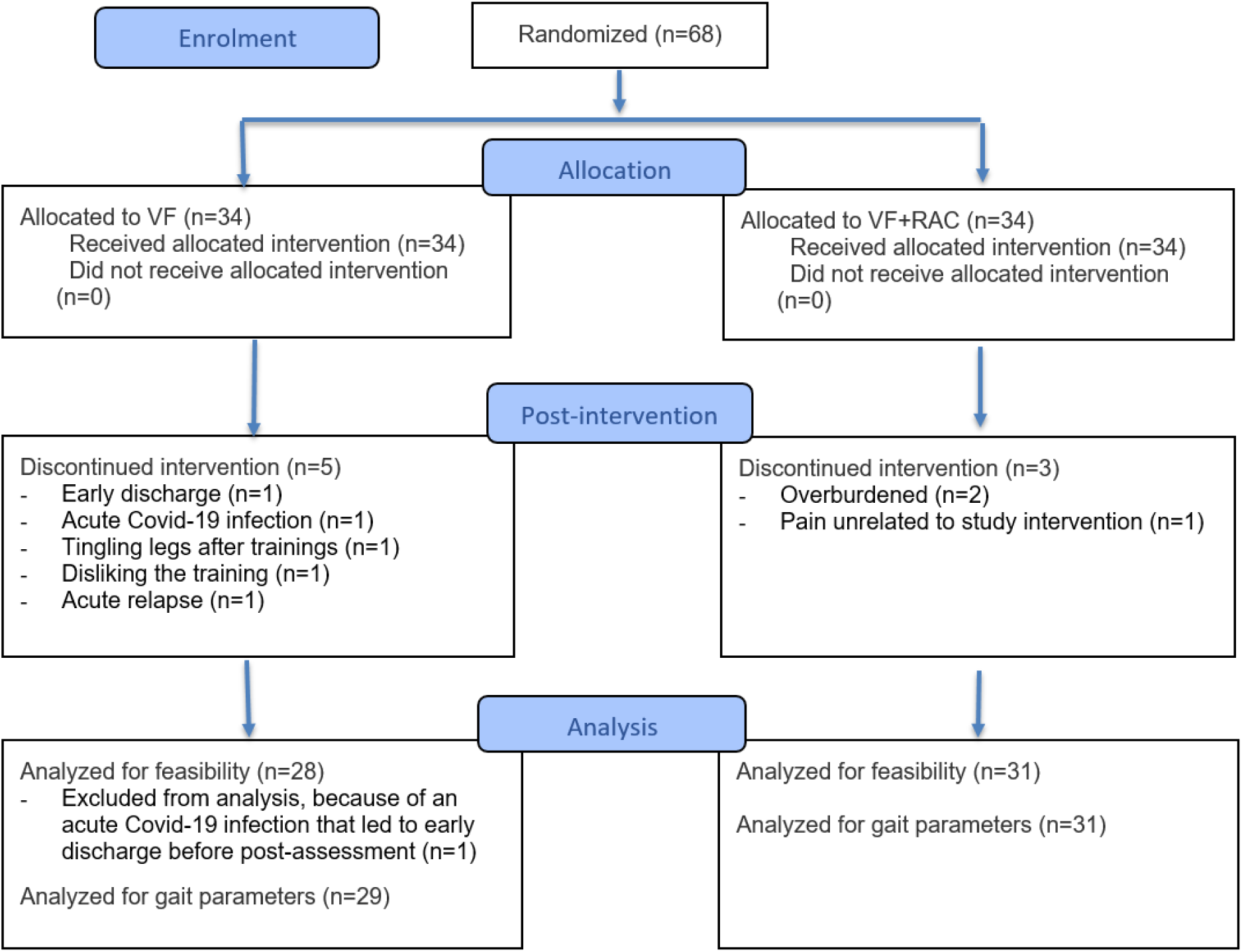
Adapted CONSORT diagram. VF=Visual feedback, VF+RAC=Visual feedback+rhythmic auditory cueing.

At baseline, participants in both groups had similar sociodemographic and clinical characteristics except for level of education (Table 1).

**Table 1.** Baseline sociodemographic and clinical characteristics.

|  | VF (n=34) | VF+RAC (n=34) | p-value |
| --- | --- | --- | --- |
| Age in years mean (*SD*, range) | 49 (9, 32 – 63) | 46.3 (11.1, 23 – 63) | .277<sup>a</sup> |
| Sex female:male | 24:10 | 22:12 | .604<sup>b</sup> |
| MS type RR:SP:PP (*n*) | 26:2:5 | 26:6:2 | .220<sup>c</sup> |
| TSD in years mean (*SD*, range) | 10.9 (11.5, 0.1 - 42) | 7.7 (8.3, 0.1 - 26.8) | .334<sup>d</sup> |
| EDSS median (*IQR*, range) | 2.5 (2.5, 1 – 6.5) | 3 (1, 1 – 6.5) | .705<sup>d</sup> |
| MSFC median (*IQR*, range) | 0.15 (0.55, -1.18 – 1.16) | -0.03 (0.9, -1.32 – 0.86) | .234<sup>d</sup> |
| Disease-modifying treatment yes:no | 19:8* | 23:7* |  |
| Education (*n*) |  |  |  |
| <9 years | 0 | 0 | **.036<sup>d</sup>** |
| 9-11 years | 0 | 4 |  |
| 12 years | 1 | 2 |  |
| 13-16 years | 14 | 16 |  |
| >16 years | 19 | 12 |  |
| Musicality (*n*) |  |  |  |
| No musical activity | 0 | 0 | .984<sup>d</sup> |
| Only listening to music | 15 | 17 |  |
| Only former musical activity | 16 | 11 |  |
| Low musical activity | 1 | 0 |  |
| Moderate musical activity | 1 | 2 |  |
| High musical activity | 1 | 4 |  |
| Total rehabilitation time in minutes<sup>+</sup> mean (*SD*, range) |  |  |  |
| Motor focus | 1549 (596, 675 – 3420) | 1460 (463, 630 – 2340) | .797<sup>a</sup> |
| Cognitive focus | 470 (416, 30 – 1995) | 399 (244, 60 – 1170) | .989<sup>d</sup> |

<sup>a</sup>Independent samples *t*-test, <sup>b</sup>Chi-squared test, <sup>c</sup>Fisher's Exact test, <sup>d</sup>Mann-Whitney *U* test, <sup>+</sup>including study interventions in "Motor focus", \**n*≠34 due to incomplete data in medical records, bold=significant difference between groups, EDSS=Expanded Disability Status Scale, MS=multiple sclerosis, MSFC=Multiple Sclerosis Functional Composite, PP=primary progressive multiple sclerosis, RR=relapsing-remitting multiple sclerosis, SP=secondary progressive multiple sclerosis, TSD=time since diagnosis, VF=Visual feedback, VF+RAC=Visual feedback+rhythmic auditory cueing.

### 3.2 Feasibility

#### 3.2.1 Adherence and compliance

Dropout reasons and time points are shown in Figure 3. Sixty of 68 participants (88%) attended at least eight of ten training sessions (the pre-specified minimum number, VF: 29/34 (85%) and VF+RAC: 31/34 (91%)) and completed the intervention period. Mean number of attended sessions in the VF-group were 9.2 (*SD*: 1.7) and 9.3 (2.0) in the VF+RAC-group (including dropouts). Of the 680 sessions initially planned, 629 (93%) (VF: 312/340 (92%), VF+RAC: 317/340 (93%)) were conducted (i.e., adherence), while 586 (86%) (VF: 294/340 (86%), VF+RAC: 292/340 (86%)) were completed as prescribed (duration of 30 minutes, i.e., compliance). The 43 sessions conducted but not completed as prescribed were stopped after 20 minutes on average in both groups. In session 1, ten (VF) and eleven (VF+RAC) participants needed at least one rest break, while this decreased to seven participants in session 10, for each group. I.e., about two thirds of participants completed the 30-minute sessions without any rest breaks.

Examination of the rate of synchrony showed that the VF+RAC-group was 73.5% (*SD*: 16.1) compliant with the metronome in session 1, improving to 87.8% (15.4) in session 10. Compliance with the music beat was 80.8% (15.3) and 91.3% (14.6), respectively. Overall, synchrony to the music was higher (see Figure 4H)). Since many participants initially demonstrated insufficient step length along with elevated cadence, increasing the metronome’s/music’s tempo was often more suitable in the beginning of the intervention (i.e., start: tempo +10%, decreasing to +5% and 0% in the following sessions if suitable).

**Figure 4.**
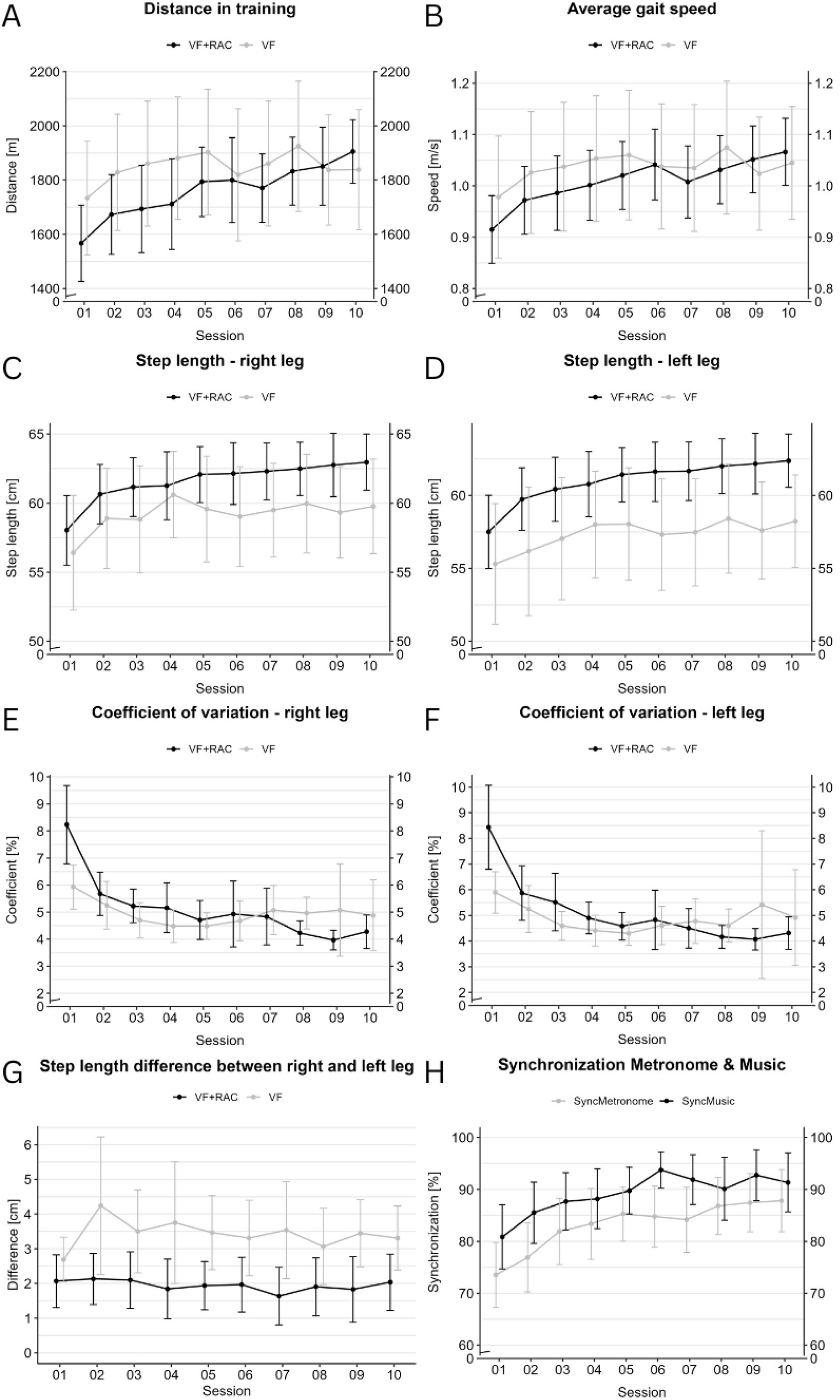
Session-by-session gait parameters and synchronization. VF=Visual feedback, VF+RAC= Visual feedback+rhythmic auditory cueing

#### 3.2.2 Safety

No SAEs occurred in either group. Thirty-eight AEs occurred in 30 of 629 sessions (VF: *n*=16, VF+RAC: *n*=22), meaning that AEs happened in 6% of all sessions (VF: 5%, VF+RAC: 7%). In both groups, AEs occurred in nine individuals, with a maximum of seven and five AEs for one individual in the VF and VF+RAC-group, respectively. The most frequently reported AE was pain (*n*=21 (55%)) (VF: *n*=7 (44%), VF+RAC: *n*=14 (64%)), followed by dizziness (*n*=9 (24%)) (VF: *n*=6 (38%), VF+RAC: *n*=3 (14%)), and tingling legs post-training (*n*=4 (11%)) (VF: *n*=2 (13%), VF+RAC: *n*=2 (9%)). Three falls occurred unrelated to a rehabilitation activity (VF: *n*=1, VF+RAC: *n*=2) and once during a VF+RAC session. In this single case the session had to be terminated, but the participant was able to continue the intervention on the following day. Risk ratios for the occurrence of AEs in individual patients and for the number of sessions with AEs showed no differences between groups (RR=1, *CI*_95%_=[0.31, 1.49] and RR=1.03, *CI*_95%_=[0.32, 1.3]).

#### 3.2.3 Acceptability

Participants’ rehabilitation goals and the perceived relevance of the study interventions to reach goals are provided in Supplement 2.1 and Supplement 2.2. There was no difference between groups regarding perceived relevance of the interventions (*p*=.571).

Also, no statistically significant group difference was observed (*p*=.579) regarding system usability (SUS), and both groups reported high usability (VF: 91.2 (median, *IQR*=15, range: 57.5-100, *n*=28), VF+RAC: 90 (10, 67.5-100, *n*=29)).

In the TEI, the VF-group scored a median 12 out of 13 points (*IQR*=2.62, range: 8-13, *n*=28) as did the VF+RAC-group (2.5, 6-13, *n*=31), indicating very good experiences with both training programs. Accordingly, no statistically significant group difference was found (*p*=.583).

Descriptive data for emotions before and after training are displayed in Supplement 2.3. On average, 77% (80% VF, 75% VF+RAC) of participants were in “good mood” or “very good mood” and 75% (75% VF, 75% VF+RAC) were “rather motivated” or “highly motivated” before the training sessions. 62% of participants (64% VF, 62% VF+RAC) were “barely exhausted” or “not exhausted at all” after the training sessions, meaning also that 38% were “a little exhausted” or “very exhausted” after the training. Lastly, 87% (89% VF, 86% VF+RAC) indicated that the training was “a little fun” or “a lot of fun”.

### 3.3 Gait parameters

Descriptive data of session 1 and 10 are presented in Table 2. Figure 4 displays session-by-session gait parameters. In session 1, no statistically significant group differences in spatiotemporal gait parameters (distance, gait speed, and step length) were observed. However, step length variability (CoV) was significantly higher in the VF+RAC-group (right *p*=.016 and left leg *p*=.019), while step length difference was significantly smaller compared to the VF-group (*p*=.021).

**Table 2.** Descriptive data for gait parameters of session 1 and 10.

| Outcome | VF | VF+RAC |
| --- | --- | --- |
| Spatiotemporal gait parameters |  |  |
| Distance (m) |  |  |
| S1 | 1733 (552), *n*=29 | 1567 (375), *n*=30 |
| S10 | 1839 (548), *n*=26 | 1905 (308), *n*=29 |
| S10-S1 (change) | **137 [43, 231], *n*=26** | **275 [193, 358], *n*=28** |
| **Gait speed (m/s)** |  |  |
| S1 | 0.98 (0.31), *n*=29 | 0.92 (0.18), *n*=30 |
| S10 | 1.05 (0.27), *n*=26 | 1.07 (0.17), *n*=29 |
| S10-S1 (change) | **0.09 [0.05, 0.13], *n*=26** | **0.13 [0.08, 0.18], *n*=28** |
| **Step length right (cm)** |  |  |
| S1 | 56.4 (10.9), *n*=29 | 58 (6.7), *n*=30 |
| S10 | 59.8 (8.5), *n*=26 | 63 (5.4), *n*=29 |
| S10-S1 (change) | **4 [1.4, 6.5], *n*=26** | **4.6 [3, 6.4], *n*=28** |
| **Step length left (cm)** |  |  |
| S1 | 55.3 (10.9), *n*=29 | 57.5 (6.7), *n*=30 |
| S10 | 58.2 (7.8), *n*=26 | 62.4 (4.8), *n*=29 |
| S10-S1 (change) | **3.4 [1, 5.7], *n*=26** | **4.6 [2.7, 6.5], *n*=28** |
| **Qualitative gait parameters** |  |  |
| **CoV right (%)** |  |  |
| S1 | 5.9 (2.1), *n*=28 | 8.2 (3.9), *n*=30 |
| S10 | 4.9 (3.2), *n*=25 | 4.3 (1.6), *n*=29 |
| S10-S1 (change) | **-1.2 [-2.3, 0.01], *n*=25** | **-3.8 [-5.4, -2.2], *n*=28** |
| **CoV left (%)** |  |  |
| S1 | 5.9 (2.1), *n*=28 | 8.4 (4.4), *n*=30 |
| S10 | 4.9 (4.5), *n*=25 | 4.3 (1.7), *n*=29 |
| S10-S1 (change) | **-1 [-2.8, 0.7], *n*=25** | **-3.9 [-5.5, -2.3], *n*=28** |
| **Step length difference (cm)** |  |  |
| S1 | 2.7 (1.7), *n*=29 | 2.1 (2), *n*=30 |
| S10 | 3.3 (2.3), *n*=26 | 2 (2.1), *n*=29 |
| S10-S1 (change) | 0.6 [-0.01, 1.2], *n*=26 | **-0.1 [-0.6, 0.3], *n*=28** |

All data indicate means and standard deviations (SD) or 95% confidence intervals [CI<sub>95%</sub>], bold=improvements, S1=session 1, S10=session 10, VF=Visual feedback, VF+RAC=Visual feedback+rhythmic auditory cueing, CoV=Coefficient of variation. Note: change scores are based on individual changes and not on group level differences between session 1 and session 10.

Mean improvements from session 1 to session 10 were observed in both groups for all gait parameters except for step length difference in the VF-group, while it remained constant in the VF+RAC-group (about 2cm). Average improvement in distance walked during a treadmill session was twice as large for the VF+RAC-group compared to the VF-group.

Figures on session-by-session developments indicated an overall higher variance in the spatiotemporal gait parameters (i.e., larger confidence intervals) in the VF-group, a somewhat steady increase in distance walked and gait speed in the VF+RAC-group compared to a plateau after session 5 in the VF-group, and overall greater step lengths in the VF+RAC-group. When excluding session 1 (see discussion), CoV remained somewhat constant in the VF-group, while for the VF+RAC a slight but continuous improvement was present. Step length difference remained around 2cm for the VF+RAC during all sessions. Asymmetry increased in the VF-group from session 1 to session 2 and then remained at around 3.5cm.

## 4 Discussion

This study aimed to investigate the feasibility of treadmill training with visual feedback or visual feedback plus rhythmic auditory cueing during inpatient rehabilitation for pwMS. Secondarily, effects on spatiotemporal and qualitative gait parameters were explored. Feasibility results showed good adherence and compliance rates, appropriate safety without SAEs and only one fall during training, and high acceptability. Both groups showed increases in distance, gait speed, and average step length. Step length variability (i.e. CoV) remained constant in the VF-group, while the VF+RAC-group slightly improved. Step length difference was around 2cm for the VF+RAC-group during all sessions and increased in the VF-group.

Exercise adherence is a prerequisite for its efficacy (Motl et al., 2023). Especially in inpatient rehabilitation settings, where study interventions are usually provided as an add-on to the usual care rehabilitation activities, it becomes an important aspect of feasibility (Wolf et al., 2023). In the present study, pwMS participated in high-frequency treadmill training (five sessions per week) and received an average total of 25h of motor rehabilitation and around 7h of cognitive rehabilitation during the intervention period (i.e., 2 weeks). Despite this dense schedule, results still showed high adherence for both groups and only slightly higher dropouts (12%) than the average in MS exercise RCTs (10%) (Motl et al., 2024). This is in line with previous studies conducted in similar inpatient rehabilitation settings reporting adherence rates at or above 90% (Wolf et al., 2023; Zimmer et al., 2018).

Even though the compliance rate (86%) was not considerably lower than adherence, and above the published average of 70% (Motl et al., 2024), overall compliance data indicated a subsample of pwMS, who were unable to fully comply. E.g., for 13 individuals, sessions had to be terminated early (VF: 7, VF+RAC: 6). Some participants needed rest breaks to complete the 30 minutes on the treadmill and 39% reported to be “a little exhausted” or even “very exhausted” after the training. This supports that inpatient rehabilitation provides a setting in which pwMS are willing to engage in a challenging training load (Wolf et al., 2024), but for some, the therapist might need to adapt the load, e.g., via an adapted session duration of 20 minutes, as this was the average time, when preliminary terminated sessions were stopped.

AEs occurred in both study groups with leg pain reported the most. The occurrence of AEs in exercise studies in MS is common. In fact, AEs are reported in 88% of studies (Learmonth et al., 2023), but the total number of AEs was considerably higher in the present study (32 vs. 0-7). We hypothesize this to be related to our strict monitoring of AEs and the difficulty of distinguishing whether AEs were directly related to the treadmill training or related to the preceding rehabilitation activities or the cumulative effect thereof. Notably, the only adverse event with a clear relation to the intervention, which is also referred to as adverse effect (Learmonth et al., 2023), was one fall in the VF+RAC-group. Moreover, 44% of total events in the VF-group, and 23% of total events in the VF+RAC-group were related to a single individual.

Regarding acceptability, there were hardly any differences between VF and VF+RAC. Both groups experienced the Gait Trainer 3.1 to have high usability, as SUS scores of 90 and above can be graded as an “A” (best grade possible) (Bangor et al., 2009). With respect to the study of Maggio and colleagues, where pwMS exercised on the same treadmill, mean SUS score in their VF+RAC-group was 86 (Maggio et al., 2021), supporting the present results.

Despite that pre-intervention rehabilitation goals were diverse and not necessarily specific to the intervention, 54 of 59 participants (92%) rated the treadmill training to be highly or at least rather relevant to achieve their individual goals. This may have contributed to the participant’s motivation to engage in the training. Three out of four participants stated to be “highly motivated” or at least “rather motivated”, regardless of group allocation and most participants (86%) in both groups reported to have had either “a little bit of fun” or “a lot of fun”, which may also have facilitated retention and compliance. Results on qualitative gait parameters could have important practical implications. Therapists should be aware that introducing RAC initially increases step length variability, as participants try to adapt to the exercise. Furthermore, finding the preferred treadmill speed also increases the CoV, because adaptation to different speeds requires adjustment of step length and cadence. Therefore, change from session 1 is not adequate to interpret this parameter. Nevertheless, comparison with published literature shows that after the first three sessions, average CoV in both groups was at or below the level reported in usual walking studies in pwMS (5.1%), but was still elevated compared to healthy adults (2%) (Socie et al., 2013). Interestingly, already in session 1, RAC consistently brought average step length difference close to that of healthy adults in the VF+RAC-group (2cm vs. 1.8cm) (Kalron, 2015). Contrarily, VF for step length alone seemed to increase asymmetry between the left and right leg, as the lowest asymmetry in the VF-group was present in the first session. Counterintuitively, VF for step length was not sufficient to facilitate a more symmetrical gait. Instead, RAC seems to be more promising, potentially via its facilitation of planning and execution of cyclical movements (Helmlinger et al., 2025). Even more when considering that the first ten minutes of every training session were spend without RAC. Our assessment of the rate of synchronization promotes the assumption that a more symmetrical gait can already be achieved when being just about 80% compliant with the beat, as less asymmetry already occurred in the first session without any further improvement even when synchronization rates increased.

For interpretation of studies already indicating positive effects of RAC on gait parameters and walking performance in pwMS (Conklyn et al., 2010; Maggio et al., 2021; Shahraki et al., 2017), larger sample sizes would have been desirable. Observations from this study support that RAC provides a benefit for walking ability, although treadmill training incorporating VF alone also yields benefits for walking distance, gait speed, and step length, which has already been shown in other neurological conditions such as stroke (Kaźmierczak et al., 2022; Kim and Oh, 2020) or older adults (Silva-Batista et al., 2023).

Several limitations must be considered. From a design perspective, it would have been more appropriate to have uninvolved researchers perform randomization and group assignment. Furthermore, criteria for successful feasibility were not defined *a priori* (Eldridge et al., 2016). Also, the (non-)eligibility rate was not monitored and could have provided valuable information for future studies. Regarding interpretation of the gait parameters, inclusion of a treadmill-only group and assessment of steady-state treadmill walking without feedback or RAC, pre- and post-intervention would have been desirable. Therefore, we were not able to directly observe the effect of the interventions on usual gait. For participants who had to terminate sessions early, parts of the metronome and/or music-based RAC were missing on that day. This might have led to a disproportionally lower stimulus of music-based RAC compared to VF and metronome-based RAC. Furthermore, there was no predetermined progression of training load in terms of treadmill speed, providing the possibility of an individualized approach, but also compromising reproducibility.

Future studies should investigate effects of VF and RAC on joint-level gait parameters, as these might be even more sensitive to early-stage gait deviations. In the future, this might be accomplished via markerless systems, making it more applicable in clinical settings (Ramari et al., 2026). Furthermore, diverging effects of treadmill training with metronome or music-based RAC should be investigated, as well as the effect of walking to one’s favorite music genre. Powering future studies *a priori* to detect clinically relevant difference in qualitative gait parameters will also be an important objective. In practice, the training duration (30 minutes) should be adapted to the participant’s walking capacity, so that training duration is at least 20 minutes with the option of adding ten minutes when feeling confident. Further, predetermined exercise progression criteria should be adopted but focus on individualized approaches that reflect the heterogeneity of pwMS in rehabilitation settings.

## 5 Conclusions

For pwMS who were eligible for participation in either intervention, VF and VF+RAC are feasible and safe training options. VF and VF+RAC are well received by participants and perceived to contribute to achievement of rehabilitation goals. Improvements in spatiotemporal gait parameters were observed in both groups, while improvements in gait variability and symmetry were more pronounced in the VF+RAC-group and therefore attributed to RAC.

## Acknowledgements

We thank all participants for their time and effort. We also thank the neuropsychological staff of the JKG for their support with screening and recruitment.

## Author contributions

Conceptualization: ME, JS, and JN, data curation: PK and ME, formal analysis: PK and FW, funding acquisition: *n*.a., investigation: PK, JS and ME, methodology: ME and JS, project administration: PK, ME and JS, resources: PK, FW, JS, JN, and ME, software: PK, supervision: PK, JS and ME, Validation: PK, visualization: PK and FW, writing – original draft: PK and FW, writing – review & editing: PK, FW, JS, JN, and ME

## Data availability

The raw data were generated at the Johanniter-Klinik Godeshöhe GmbH. The datasets used and/or analyzed during the current study are available from the corresponding author on reasonable request.

