## Supplement 2 for "Treadmill training with rhythmic auditory cueing and/or visual feedback for persons with Multiple Sclerosis: feasibility and effects on gait parameters in a clinical randomized controlled trial" (from another version of this preprint: version 1, 2026-06-22)

**Table S2.1**. Participants’ rehabilitation goals pre-intervention.

| Domain | Category | VF+RAC (n=34) | VF (n=34) |
| --- | --- | --- | --- |
| Physical functions | Increasing walking distance/endurance | 17 | 11 |
|  | Improving walking safety/ability | 9 | 9 |
|  | Improving balance | 6 | 6 |
|  | Improving physical strength | 5 | 7 |
|  | Improving coordination | 0 | 3 |
|  | Retaining current health/fitness level or increasing overall physical fitness level | 7 | 8 |
|  | Reduction of body weight | 3 | 2 |
|  | Improvement of fine motor skills | 3 | 4 |
| Cognitive/psychological | Improvement of cognitive functions | 10 | 8 |
|  | Reduction of stress levels/improving resiliance/relaxation | 7 | 7 |
|  | Disease management or health education | 3 | 10 |
|  | Improving mood/reducing depression | 2 | 6 |
| Participation | Regaining or improving the ability to work | 10 | 8 |
|  | Being able to participate in sports again | 4 | 2 |
|  | Improving other social functions (excluding sports) | 1 | 3 |
| Disease-related goals | Fatigue management/fatigue reduction (also increasing energy levels) | 12 | 7 |
|  | Reduction of bodily pain or headaches | 5 | 1 |
|  | Improving speech problems | 2 | 0 |
|  | Improvement of vision | 2 | 0 |
|  | Improvement of tingling paresthesia | 1 | 2 |
|  | Other | 6 | 5 |

VF= Visual feedback, VF+RAC=Visual feedback+rhythmic auditory cueing, `other` goals were unspecific or only mentioned once, e.g. urological treatment, treatment of dizziness, etc.

**Table S2.2**. Perceived relevance of the study interventions to achieve individual rehabilitation goals.

| Relevance | VF (n) |  | VF+RAC (n) |
| --- | --- | --- | --- |
| Not relevant at all | 1 |  | 0 |
| Less relevant | 2 |  | 2 |
| Rather relevant | 11 |  | 9 |
| Highly relevant | 14 |  | 20 |

VF=Visual feedback, VF+RAC= Visual feedback+rhythmic auditory cueing

**Table S2.3**. Descriptive data for emotion before and after training

| Outcome | VF | | | | | | | | | | VF+RAC | | | | | | | | | |
| --- | --- | --- | --- | --- | --- | --- | --- | --- | --- | --- | --- | --- | --- | --- | --- | --- | --- | --- | --- | --- |
|  | S1 | S2 | S3 | S4 | S5 | S6 | S7 | S8 | S9 | S10 | S1 | S2 | S3 | S4 | S5 | S6 | S7 | S8 | S9 | S10 |
| **Mood** |  |  |  |  |  |  |  |  |  |  |  |  |  |  |  |  |  |  |  |  |
| Bad mood | 1 | 1 | 3 | 1 | 2 | 2 | 1 | 2 | 1 | 2 | 0 | 2 | 2 | 0 | 3 | 2 | 1 | 2 | 2 | 2 |
| Not really in a good mood | 5 | 9 | 1 | 6 | 4 | 7 | 4 | 5 | 6 | 1 | 8 | 6 | 7 | 10 | 5 | 7 | 7 | 7 | 3 | 2 |
| Good mood | 23 | 17 | 24 | 20 | 19 | 17 | 20 | 16 | 10 | 18 | 25 | 23 | 22 | 17 | 21 | 18 | 17 | 14 | 17 | 23 |
| Very good mood | 5 | 7 | 6 | 7 | 8 | 5 | 5 | 6 | 10 | 5 | 1 | 3 | 2 | 5 | 2 | 4 | 6 | 8 | 8 | 3 |
| n | 34 | 34 | 34 | 34 | 33 | 31 | 30 | 29 | 27 | 26 | 34 | 34 | 33 | 32 | 31 | 31 | 31 | 31 | 30 | 30 |
| **Motivation** |  |  |  |  |  |  |  |  |  |  |  |  |  |  |  |  |  |  |  |  |
| Not at all motivated | 1 | 1 | 0 | 2 | 1 | 2 | 0 | 3 | 1 | 1 | 2 | 2 | 1 | 2 | 1 | 2 | 1 | 2 | 3 | 2 |
| Rather not motivated | 7 | 6 | 8 | 7 | 6 | 9 | 11 | 3 | 7 | 2 | 4 | 5 | 7 | 8 | 6 | 8 | 9 | 5 | 4 | 5 |
| Rather motivated | 9 | 16 | 15 | 16 | 17 | 12 | 8 | 13 | 8 | 14 | 16 | 19 | 15 | 14 | 16 | 15 | 13 | 14 | 14 | 15 |
| Highly motivated | 17 | 11 | 11 | 9 | 9 | 8 | 11 | 10 | 11 | 9 | 12 | 8 | 10 | 8 | 8 | 6 | 8 | 10 | 9 | 8 |
| n | 34 | 34 | 34 | 34 | 33 | 31 | 30 | 29 | 27 | 26 | 34 | 34 | 33 | 32 | 31 | 31 | 31 | 31 | 30 | 30 |
| **Exhaustion** |  |  |  |  |  |  |  |  |  |  |  |  |  |  |  |  |  |  |  |  |
| Not exhausted at all | 9 | 8 | 4 | 6 | 6 | 6 | 8 | 6 | 6 | 6 | 7 | 12 | 8 | 7 | 6 | 7 | 5 | 8 | 9 | 7 |
| A little exhausted | 14 | 14 | 17 | 15 | 15 | 13 | 13 | 13 | 9 | 11 | 15 | 6 | 12 | 13 | 14 | 13 | 10 | 8 | 10 | 15 |
| Barely exhausted | 8 | 9 | 10 | 9 | 9 | 8 | 6 | 6 | 5 | 5 | 10 | 14 | 11 | 10 | 9 | 7 | 12 | 11 | 5 | 10 |
| Completely exhausted | 3 | 3 | 3 | 4 | 3 | 4 | 3 | 4 | 7 | 4 | 2 | 1 | 1 | 1 | 2 | 4 | 4 | 3 | 6 | 2 |
| n | 34 | 34 | 34 | 34 | 33 | 31 | 30 | 29 | 27 | 26 | 34 | 33 | 32 | 31 | 31 | 31 | 31 | 30 | 30 | 34 |
| **Fun** |  |  |  |  |  |  |  |  |  |  |  |  |  |  |  |  |  |  |  |  |
| No fun at all | 0 | 0 | 1 | 2 | 1 | 1 | 2 | 2 | 2 | 2 | 2 | 1 | 2 | 2 | 2 | 2 | 2 | 2 | 2 | 3 |
| Barely any fun | 3 | 3 | 2 | 2 | 0 | 2 | 2 | 2 | 1 | 4 | 2 | 3 | 4 | 2 | 3 | 2 | 1 | 2 | 4 | 2 |
| A little fun | 14 | 15 | 14 | 13 | 15 | 13 | 10 | 9 | 10 | 5 | 15 | 19 | 15 | 15 | 14 | 17 | 16 | 12 | 12 | 13 |
| A lot of fun | 17 | 16 | 17 | 17 | 17 | 15 | 16 | 16 | 14 | 15 | 15 | 11 | 12 | 13 | 12 | 10 | 12 | 15 | 12 | 12 |
| n | 34 | 34 | 34 | 34 | 33 | 31 | 30 | 29 | 27 | 26 | 34 | 34 | 33 | 32 | 31 | 31 | 31 | 31 | 30 | 30 |

S=Session, VF=Visual feedback, VF+RAC=Visual feedback+rhythmic auditory cueing
